# Phylogeography and transmission of *M. tuberculosis* spanning prisons and surrounding communities in Paraguay

**DOI:** 10.1101/2022.08.23.22279039

**Authors:** Gladys Estigarribia Sanabria, Guillermo Sequera, Sarita Aguirre, Julieta Méndez, Paulo César Pereira dos Santos, Natalie Weiler Gustafson, Analía Ortiz, Cynthia Cespedes, Gloria Martínez, Alberto L. García-Basteiro, Jason R. Andrews, Julio Croda, Katharine S. Walter

## Abstract

Recent rises in incident tuberculosis (TB) cases in Paraguay and the increasing concentration of TB within prisons highlight the urgency of targeting strategies to interrupt transmission and prevent new infections. However, whether specific cities or carceral institutions play a disproportionate role in transmission remains unknown. We conducted prospective genomic surveillance, sequencing 471 *M. tuberculosis* genomes, from inside and outside prisons in Paraguay’s two largest urban areas, Asunción and Ciudad del Este, from 2016 to 2021. We found genomic evidence of frequent recent transmission within prisons and transmission linkages spanning prisons and surrounding populations. We identified a signal of frequent *M. tuberculosis* spread between urban areas and marked recent population size expansion of the three largest genomic transmission clusters. Together, our findings highlight the urgency of strengthening TB control programs to reduce transmission risk within prisons, where, in Paraguay, incidence was 70 times that outside prisons in 2021.

**Financing agencies:** National Institutes of Health grants R01 AI130058 (JRA) and R01 AI149620 (JRA and JC). Paraguay National Commission of Science and Technology grant CONACYT PIN 15-705 (GES and GS).

## Introduction

Despite significant tuberculosis (TB) control efforts, the incidence rate of TB has declined only slowly in the World Health Organization Region of the Americas, and, alarmingly, has stagnated since 2014^1^. The COVID-19 pandemic disrupted access to healthcare—including critical TB diagnostic and treatment programs—compounding the burden of TB and reversing decades of progress in TB control^1^.

New approaches to limit transmission are urgently needed in Paraguay, where TB control is chronically underfunded^1^ and where TB incidence was 48 (41-56) per 100,000 people in 2020, higher than the mean incidence rate across the region^1^. More than a quarter of the country’s population lives below the national poverty line^2^ and are at heightened risk of TB infection and mortality. Further, recent dramatic increases in incarceration^3,4^ put a rapidly growing population at high risk of infection and disease^5–7^. To guide interventions in Paraguay, there is a critical need to identify the populations at greatest risk of infection and locations and institutions where transmission most frequently occurs^8^.

Whole genome sequencing of *M. tuberculosis* has been powerfully applied to characterize recent transmission dynamics. Genomic approaches have dated introductions of *M. tuberculosis* and reconstructed patterns of historic spread across Central and South America^9,10^, estimated the contribution of recent transmission to incident TB cases^11^, reconstructed the emergence of resistance-associated mutations^12^, and inferred likely individual-level transmission events^13^. In Brazil^14^ and Georgia^13^, for example, genomic approaches identified frequent transmission within prisons as well as evidence of spillover from prisons to surrounding communities. A single *M. tuberculosis* molecular study from Paraguay^15^ on strains collected in 2003 reported that *M. tuberculosis* spoligotypes found across South America including the Latin-American (LAM) and Haarlem lineages were also common in Paraguay^15,16^.

Genomic approaches have not been applied to address major gaps in our understanding of TB transmission in Paraguay. First, the conditions of incarceration put people at high risk of many infectious diseases and globally, over the past twenty years, the incarcerated population in Central and South America has grown by 206%, the greatest increase in the world^17^. Escalating incarceration rates have been paralleled by an increasing concentration of notified TB among incarcerated individuals^6^. Yet the role of prison environments on TB transmission both inside and beyond prisons, as sources of broader infection, has not yet been described in Paraguay. Second, while incidence of TB is heterogeneous across the country^15^, it remains unknown whether specific cities or regions function as hotspots, fueling transmission elsewhere. Finally, due to limited surveillance infrastructure, the prevalence of drug-resistance and multi-drug-resistance has not yet been systematically measured^18–20^. Only 56% of bacteriologically confirmed new cases of pulmonary TB were tested for rifampicin resistance in 2020^1^.

To characterize transmission dynamics and circulating diversity of *M. tuberculosis* in Paraguay, we conducted prospective genomic surveillance across the country from 2016 to 2021, including surveillance within and outside prisons, generating a genomic resource for continued surveillance in Paraguay. We estimated the role of likely recent *M. tuberculosis* transmission within prisons, the relatedness of prison and community transmission, and the frequent movement of *M. tuberculosis* between Paraguay’s urban centers.

## Methods

### Prospective population surveillance

We conducted population-based genomic surveillance in three of Paraguay’s departments: Central, Distrito Capital, and Alto Paraná, which together comprise approximately half (3,392,429 people) of the country’s 2021 population of 7.4 million. Sputum samples are routinely collected from all individuals presenting with symptoms of TB at primary health clinics and sent to the National Program for Tuberculosis Control (NPTC) of Paraguay reference laboratory for microbial diagnostics including culture and smear microscopy.

Study recruitment was done by study staff who visited patients at home and in prisons at the time patients began treatment (Directly Observed Therapy, DOT). At this time, the standard National TB Control Program questionnaire was conducted and patients who chose to enroll provided written consent for sequencing residual mycobacterial cultures for culture-positive samples. Study staff also collected additional demographic, clinical, residential, and epidemiolocal data, including information on history of prior or current incarceration with a structured questionnaire. The study was approved by the ethics committee of the Central Laboratory of Public Health of the Ministry of Health and Social Welfare of Paraguay (International Certification FWA N° FWAOOO20088) with code CEI-LCSP 91/010217.

### Laboratory and sequencing methods

Sputum samples were cultured in the Ogawa-Kudoh Method described previously^21,22^. Cultures were incubated at 37ºC and observed for growth twice a week for 60 days. *M. tuberculosis* DNA was extracted using Cetyltrimethyl ammonium bromide (CTAB) method^23^.

Sequencing was conducted at the Laboratorio Central de Salud Pública (LCSP), Paraguay Ministry of Health; Centro para el Desarrollo a la investigación Científica (CEDIC), Paraguay; and the Translational Genomics Research Institute (TGen), Arizona, US. DNA sequencing libraries were prepared with the Illumina DNA Prep library kit and sequenced on an Illumina MiSeq in Paraguay and an Illumina NextSeq (2 × 151-bp), at TGen. Raw sequence reads for samples passing filters are available at the Sequence Read Archive (PRJNA870648).

### Variant identification

We identified *M. tuberculosis* genomic variation from whole genome sequence data with a previously described pipeline available at https://github.com/ksw9/mtb_pipeline^24^. Briefly, we trimmed low-quality bases (Phred-scaled base quality < 20) and removed adapters with Trim Galore (stringency=3)^25^. We used CutAdapt to further filter reads (--nextseq-trim=20 --minimum-length=20 --pair-filter=any)^26^.To exclude potential contamination, we used Kraken2 to taxonomically classify reads and removed reads that were not assigned to the *Mycobacterium* genus or that were assigned to a *Mycobacterium* species other than *M. tuberculosis*^27^. We mapped reads with bwa v. 0.7.15 (bwa mem)^28^ to the H37Rv reference genome (NCBI Accession: NC_000962.3) and removed duplicates with sambamba^29^. We called variants with GATK 4.1 HaplotypeCaller^30^, setting sample ploidy to 1, and GenotypeGVCFs, including non-variant sites in output VCF files. We included variant sites with a minimum depth of 10X and a minimum variant quality score 40 and constructed consensus sequences with bcftools consensus^31^, excluding indels. We excluded SNPs in previously defined repetitive regions (PPE and PE-PGRS genes, phages, insertion sequences and repeats longer than 50 bp)^32^. We identified sub-lineage and evidence of mixed infection with TBProfiler v.4.2.0 ^33,34^. We additionally used TBProfiler with the TBDB repository (https://github.com/jodyphelan/tbdb) to identify resistance-associated mutations^33,34^. We described overall patterns of genomic drug-resistance including mutations that are included in the World Health Organisation catalogue as independently associated with resistance^35^.

### Phylogenetic and Bayesian evolutionary analysis

We constructed full-length consensus FASTA sequences from VCF files, setting missing genotypes to missing, and used SNP-sites to extract a multiple alignment of internal variant sites only^36^. We used the R package *ape* to measure pairwise differences between samples (pairwise.deletion=TRUE)^37^. We selected a best fit substitution model with ModelFinder^38^, implemented in IQ-TREE multicore version 2.2.0^39^, evaluating all models that included an ascertainment bias correction for the use of an alignment of SNPs only. The best fit model according to Bayesian Information Criterion was K3Pu+F+ASC+R5, a three substitution types model with unequal base frequencies, an ascertainment bias correction, and a FreeRate model of rate heterogeneity across sites, including four categories. We then fit a maximum likelihood tree with IQ-TREE, with 1000 ultrafast bootstrap replicates^39,40^.

Genomic clustering is often used as a proxy measure of recent *M. tuberculosis* transmission; isolates that are more closely genetically related are hypothesized to be more likely linked through recent transmission rather than travel-associated importation or re-activation of genetically distinct latent infections.^41,42^ We applied a commonly used genetic distance thresholds of 12- and 5- or fewer SNPs to identify genomic clusters^43,44^.

To investigate transmission patterns in the three largest genomic clusters more closely, we fit timed Bayesian trees to multiple sequence alignments with BEAST 2.6.2^45^, using TB notification dates to calibrate tips. Because of the short sampling timeframe of our data, we fixed the substitution rate to 1×10^−7^ mutations/site/year, as previously described^46^, and consistent with previous estimates for the *M. tuberculosis* lineage 4 substitution rate^47^. To examine population dynamics in the three largest clusters, we used a Coalescent Bayesian Skyline model^48^ with 5 dimensions, allowing the effective population size to change 4 times over the tree. We additionally fit a Bayesian tree to sublineage 4.2.1.1 samples using a constant population size, fixed substitution rate model. Markov chain Monte Carlo chains were run for 200 million iterations, or longer, if required for convergence, excluding 10% of samples as burn-in. We used *treeannotator* to produce maximum clade credibility trees. We used the R package *beautier* to construct XML files^49^ and corrected XML files for the number of constant positions in SNP alignments. We visualized phylogenetic trees with the R package *ggtree*^50,51^.

We calculated time-scaled haplotype density from a matrix of pairwise SNP distances with the R package *thd* as previously described^52^ and compared time-scaled haplotype density between individuals who were never, formerly, or currently incarcerated with t-tests. We set the *M. tuberculosis* substitution rate to 1×10^−7^ substitutions per site per year and included an effective genome length of 3,916,441 basepairs (the length of the reference genome minus the PE/PPE regions excluded from variant calling) and used a short (20 year) and long (50 year) epidemic timescale. We compared time-scaled haplotype density by incarceration status with t-tests and used analysis of variance to test for the independent effect of incarceration status after controlling for *M. tuberculosis* population structure, as previously described^52^.

We conducted discrete ancestral state reconstruction for sampling location with the R package *ape* for the three largest sub-lineages in our collection^37^. We restricted samples to those from Asunción and Ciudad del Este because of the small sample size outside those cities. We compared symmetric and asymmetric rates models fit with the R package *diversitree* (make.mk2) and compared model fits with analysis of variance^53^. We used stochastic character mapping^54^ in the R package *diversitree*^53^ to sample 500 location histories for each sublineage tree; we summarized these as the number of average movements between cities over the tree.

## Results

### Population-based genomic surveillance

From 2016 to 2021, 16,734 TB cases were notified in Paraguay, with the majority of cases (60%; 10,095/16,734) occurring in the urban departments Central, Distrito Capital, and Alto Paraná, where we conducted prospective genomic surveillance (Fig. 1a). In 2021, the TB notification rate was 70 times higher in prisons than outside (3,378 cases per 100,000 in prisons/49 cases per 100,000 in the general population) (Fig. 1b). Therefore, we included focused genomic surveillance in the two largest prisons in the country, Tacumbú Prison and the Prison of Ciudad del Este, which together hold 36% (4,950/13,821) of Paraguay’s incarcerated population, notification rates are 2,000 and 3,500 per 100,000 people, respectively.

**Figure 1.**
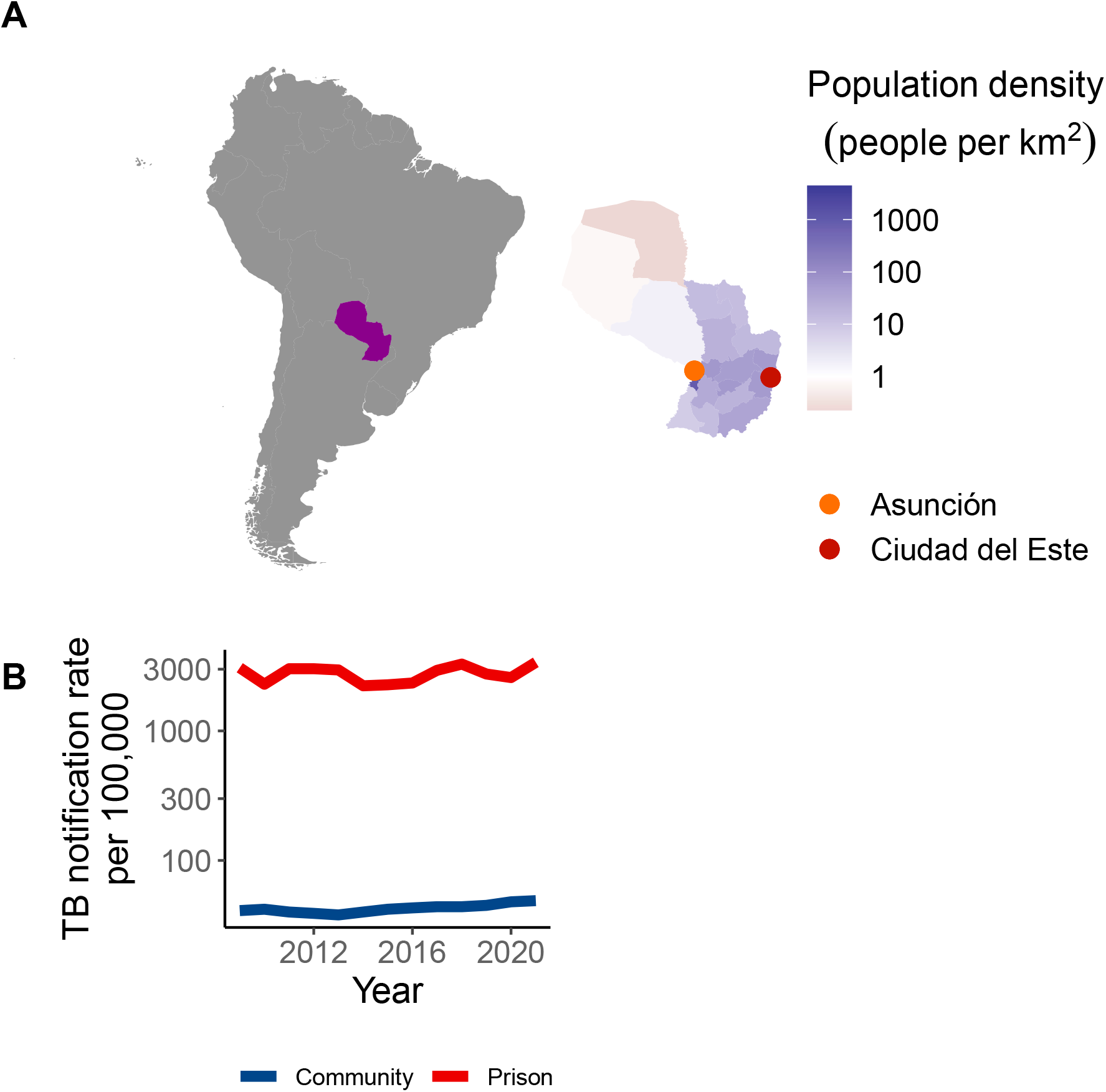
Genomic surveillance within and outside prisons in Paraguay’s urban centers. a) Map of South America, with Paraguay highlighted, and as an inset. Paraguay’s departments are colored by population size density and points indicate the two largest urban centers in Paraguay, where we conducted focused genomic surveillance. (b) Notification rate of TB per 100,000 people in prisons and in the general population from 2009 to 2020.

Whole genome sequences (WGS) for a total of 532 isolates met our coverage and quality criteria (Methods), including 488 from unique TB notifications. Of the samples passing filters, 158 were from individuals diagnosed with TB while in prison and 330 were from people diagnosed in the community. TB isolates were collected in Asunción (274/488) and in Ciudad del Este (214/488). We excluded 17 isolates with evidence of mixed infection with more than one sub-lineage detected, resulting in 471 *M. tuberculosis* isolates for following analyses.

### Genotypic resistance

The majority, 96% (454/471) of sampled *M. tuberculosis*, were drug sensitive; 3% (15/471) were resistant to at least one drug; and 0.42% (2/471) were multi-drug resistant, resistant to both isoniazid and rifampin. Resistance was not associated with sub-lineage (*X*^*2*^ (11) =7.7, p=0.74). We identified three unique isoniazid resistance-conferring mutations on the genes *fabG1, katG*, or both among the 10 isolates with any isoniazid resistance; the three rifampicin-conferring mutations in *rpoB* (two on multi-drug resistant isolates) were unique (Fig. 2).

**Figure 2.**
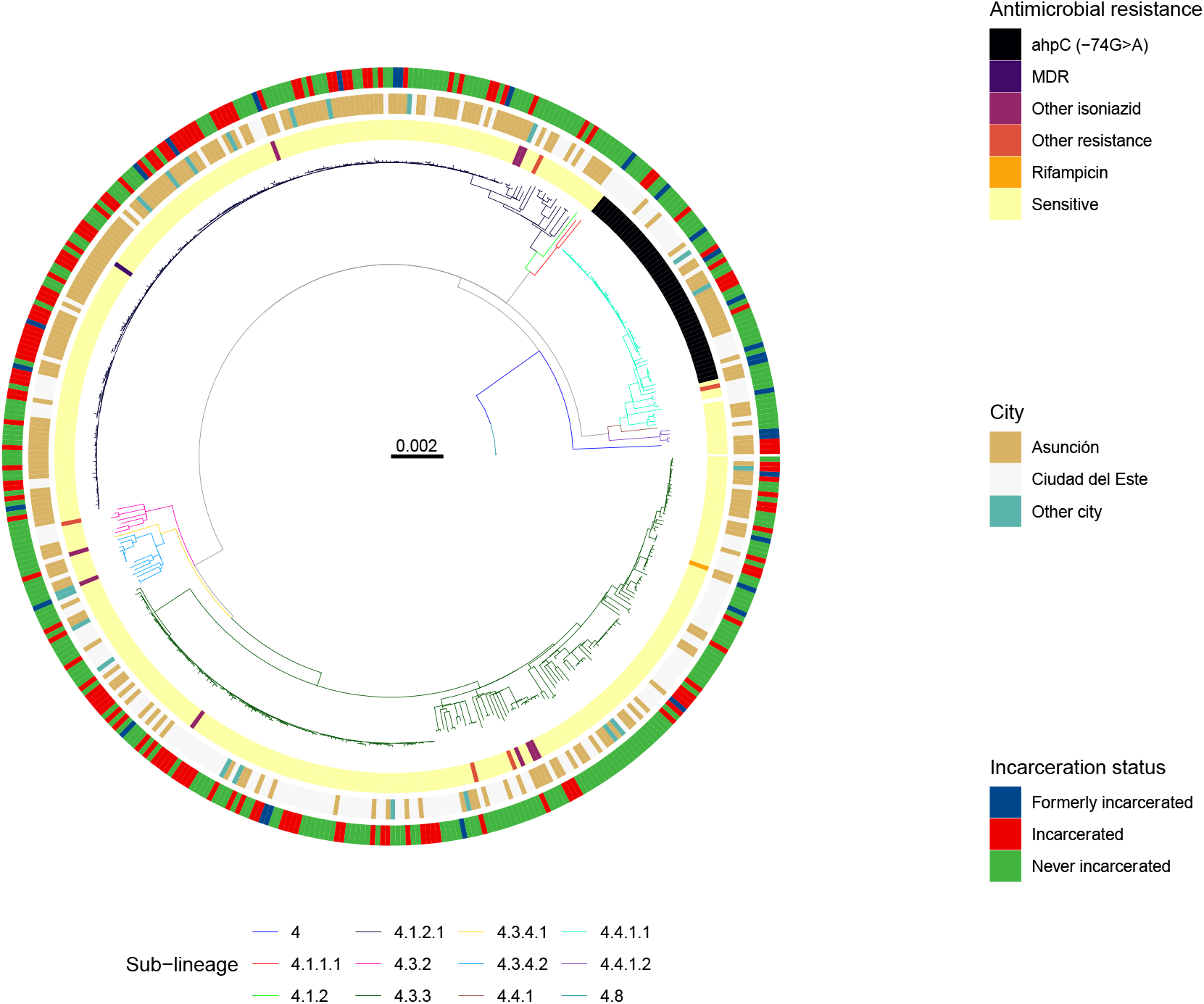
*M. tuberculosis* isolates from incarcerated and non-incarcerated people are closely related across Paraguay. A maximum likelihood phylogeny of 471 tuberculosis isolates from Lineage 4 inferred from a multiple sequence alignment of 9,966 SNPs and rooted on three sub-lineage 4.8 samples from this study. Branch lengths are in units of substitutions per site. Branches are colored by sub-lineage. From the inside, rings are colored by antimicrobial resistance; city of sampling; and incarceration status at the time of TB notification. Other isoniazid includes isoniazid mono-resistant isolates without an *ahpC* promoter mutation. Rifampicin indicates rifampicin mono-resistant isolates. Other resistance includes isolates with mutations associated with resistance to pyrazinamide, streptomycin, or fluoroquinolones and not isoniazid or rifampicin.

### Stable genomic diversity of *M. tuberculosis* in Paraguay

After excluding mixed infections, all *M. tuberculosis* isolates were from *M. tuberculosis* lineage 4. The single mixed infection was co-infected with both lineages 1 and 4. Samples predominantly fell into four sub-lineages: 4.3.3/LAM (42.5%; 200/471), 4.1.2 /Haarlem (38.2%; 180/471), 4.4.1 /S (12.3%; 58/471), and 4.3.4/LAM (3.2%; 15/471) (Fig. 2). The distribution of these sublineages was stable and did not change significantly from a collection of 173 *M. tuberculosis* isolates collected in 2003^15^ (Fig. S1).

### Geographic structure despite frequent migration across *M. tuberculosis* sub-lineages

We found a strong pattern of geographic structure (Fig. 3), with sub-lineage 4.1.2.1 dominant in Asunción (54.1%, 142/262 samples) and sub-lineage 4.3.3 dominant in Ciudad del Este (60.8%, 127/209) (*χ*2(11)=81, *p*<0.001) (Fig. 3). While we observed geographically distinct patterns of *M. tuberculosis* diversity in Asunción and Ciudad del Este, reconstruction of the ancestral locations for the three most prevalent sub-lineages revealed frequent movement of *M. tuberculosis*. Ancestors of sampled sub-lineage 4.1.2.1 isolates likely moved between the two major cities at least 150 times, since the most recent common ancestor (MCRA) of the sub-lineage in Paraguay; sub-lineages 4.4.1.1 and 4.3.3 similarly moved more than 100 times between the urban centers (Fig. 3).

**Figure 3.**
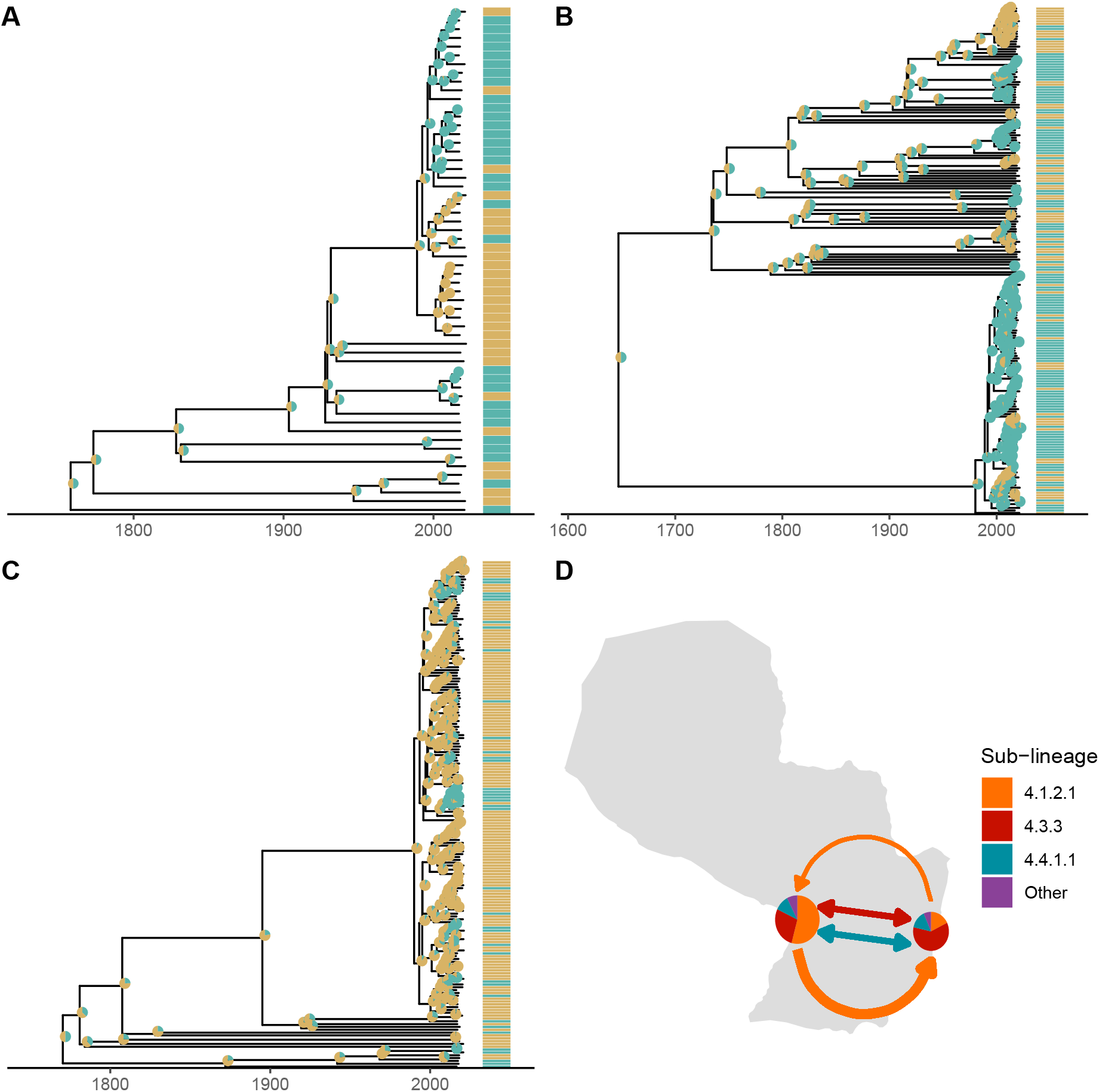
Frequent gene flow of *M. tuberculosis* connects Paraguay’s major urban centers. (a-c) We used discrete ancestral state reconstruction to reconstruct migration between the two cities for the three dominant sub-lineages in our sample; lineages 4.1.2.1, 4.3.3, and 4.4.1.1. Bayesian maximum clade credibility trees of samples in the three dominant sub-lineages with tip points colored by city of sampling and pie charts at nodes indicating the inferred ancestral location. Branch lengths are in years and grey bars indicate 95% high posterior density estimates of node date. (d) Map of Paraguay with pie charts indicating the genomic diversity sampled in Asunción (at Paraguay’s western border) and Ciudad del Este (eastern border). Arrows are colored by sub-lineage and are weighted by the relative rate of migration between cities. Bi-directional arrows indicate equal rates of migration in each direction.

To test whether Asunción and Ciudad del Este served as sources for *M. tuberculosis*, exporting infection elsewhere, we compared rates of arrival and export of each sub-lineage. Sub-lineage 4.1.2.1 moved more frequently Asunción to Ciudad del Este (mean: 75 transitions) compared to vice versa (mean: 70 transitions), and a model for asymmetric rates was supported (*χ*2(2)=4.1, *p*=0.04) (Fig. 3). Both sub-lineages 4.4.1.1 (with the prevalent *ahpC* mutation) (*χ*2(2)=0.16, *p*=0.69) and 4.3.3 (*χ*2(2)=0.56, *p*=0.46) had similar rates of migration to and from Ciudad del Este to Asunción. Despite the geographic structure observed, there was not sufficient signal to infer a likely geographic source for any of the dominant sub-lineages.

### *M. tuberculosis* genomic clusters span prisons and the general population

We next explored evidence of recent *M. tuberculosis* transmission in Paraguay, including relatedness between *M. tuberculosis* infections within and outside of prisons. In a maximum likelihood phylogeny (Fig. 2), *M. tuberculosis* isolates sampled from incarcerated and non-incarcerated people are distributed across the tree and did not form distinct clades, indicating recent shared evolutionary history of isolates sampled from prisons and the community. However, sub-lineage was associated with incarceration status (*χ*2(22)=52.3, *p*<0.001).

TB notifications were often attributable to recent transmission, with 78% percent (369/471) of all isolates falling within 26 genomic clusters (each including 2 to 159 isolates) defined by a 12-SNP threshold^42^. Isolates from incarcerated people were more frequently clustered (92.6%, 138/149), than those from formerly incarcerated (71.0%, 22/31, *χ*2(1)=10.1, *p*=0.001) or never incarcerated people (71.8%, 209/291; *χ*2(1)=24.2, p <0.0001), likely reflecting more recent transmission within prisons. We found a similar trend with a stricter threshold of 5 SNPs, with 45.4% (214/471) of all isolates in genomic transmission clusters, including 58.3% (87/149) isolates from incarcerated individuals, 45.2% (14/31) from those formerly incarcerated, and 38.8% (113/291) from never incarcerated individuals.

We predicted that if prison and community-associated epidemics were distinct, isolates from the community would be most closely related to and cluster with other isolates from the community and vice versa. Approximately half (48.0%; 12/25) of genomic clusters including people with no incarceration history also included individuals with a recent history of incarceration. The consequence is that 85.2% (178/209) of individuals with evidence of recent transmission and no recent incarceration were within transmission clusters including individuals with prior incarceration.

We additionally quantified *M. tuberculosis* recent transmission with time-scaled haplotype diversity, a measure of the centrality of a single tip isolate to all other isolates on the tree^52^. Individuals who were incarcerated at the time of TB notification had a higher time-scaled haplotype index for a short epidemic timescale (median: 0.59, IQR: 0.24-0.72) than did formerly (median: 0.18, IQR: -0.37-0.71; t(36)=1.7, p =0.03) or never incarcerated individuals (median: 0.20, IQR: -0.71-0.66; t(360)=5.9, p<0.001) (Fig. S3). This finding was consistent across epidemic timescales considered (Fig. S3). After adjusting for population structure, we found that incarceration status was significantly associated with time-scaled haplotype diversity (one-way ANOVA: F(2,85)=85, p<0.001), evidence that the association was independent of TB lineage.

### Recent growth in *M. tuberculosis* transmission clusters

We additionally reconstructed population size dynamics of the three largest genomic clusters with a Bayesian coalescent population size model. The three largest genomic clusters, including 159, 91, and 15 samples, increased in effective population size by 200, 90, and 40-fold respectively. Cluster growth was relatively recent, with cluster most recent common ancestors (MRCA) occurring in 1998 (95% HDI: 1994-2001), 1996 (95% HDI: 1991-2000), and 1998 (95% HDI: 1992-2003) respectively, to 2021, when the most recent samples were collected (Fig. 4).

**Figure 4.**
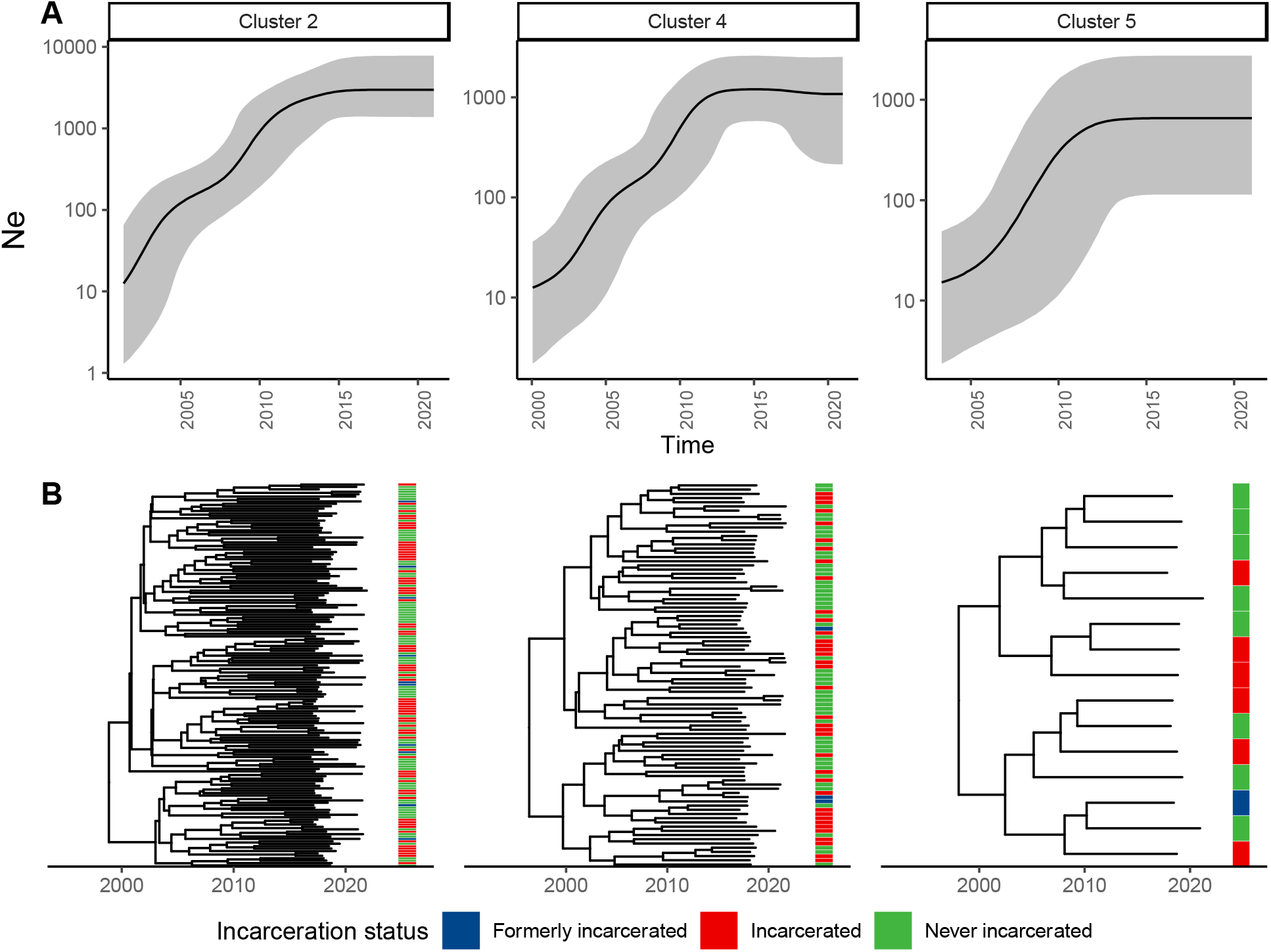
Genomic transmission clusters spanning prisons and neighboring communities have recently expanded. (a) Effective population size (*Ne*) estimates for the three largest genomic clusters in our sample over time. Black lines indicate *Ne* inferred in a Bayesian Skyline Coalescent model and grey shading indicates 95% high posterior density estimates. (b) Median clade credibility trees inferred from the Bayesian Skyline Coalescent model. Branch lengths are in years and grey bars indicate 95% high posterior density estimates of node date. The heatmap to the right of the phylogeny indicates patient incarceration status at the time of TB notification.

### Emergence of a putative resistance-associated *ahpC* promoter mutation

Eleven percent of samples (50/471) shared a mutation in *ahpC* promoter (G>A, 74 bases upstream of the 5’ start codon), previously considered a location for compensatory mutations co-occurring with *katG* mutations in isoniazid resistant isolates^35,55^. While *ahpC* promoter mutations are not included as an independent resistance-conferring mutation in the WHO resistance catalogue^35^, in our collection, *ahpC* mutations occurred on otherwise susceptible genomic background within sub-lineage 4.4.1.1. The *ahpC* mutation occurred in a monophyletic clade of 49 samples in sublineage 4.4.1.1 (Fig. 5), which shared a most recent common ancestor in 1903 (95% HDI: 1888-1916), likely reflecting a single emergence event. Among the basal group of nine samples without a fixed *ahpC* promoter locus (*ahpC*-74) mutation, one sample was polymorphic, with 16% (13/79) of reads representing the *ahpC* mutation. Among the samples sharing the *ahpC* mutation, a single isolate had a co-occurring rifampicin resistance-conferring mutation in *rpoB* (His445Leu) (Fig. 5).

**Figure 5.**
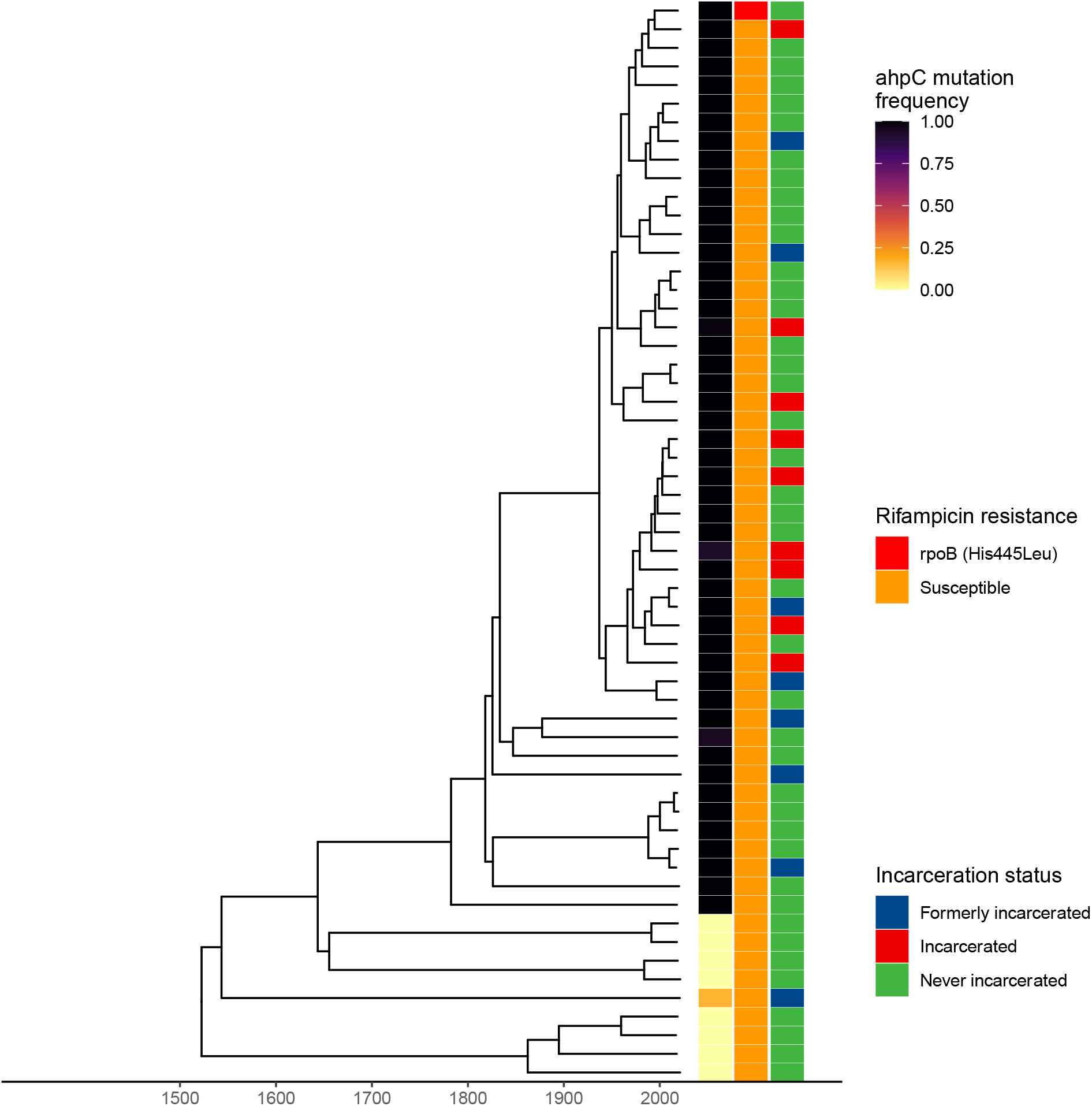
Emergence of a putative resistance-associated *ahpC* promoter mutation. Time-scaled Bayesian maximum clade credibility tree for 58 samples in sublineage 4.4.1.1. Branch lengths are in years and grey bars indicate 95% high posterior density estimates of node date. The heatmap to the right of the phylogeny indicates patient incarceration status at the time of TB notification, *ahpC* mutation frequency within an individual’s infection, and the occurrence of a rifampicin resistance-conferring mutation (*rpoB* His445Leu).

We tested whether the success of the *ahpC* mutation in the absence of a *katG* mutation (i.e. outside of a compensatory context) we observed in Paraguay could be explained by an increase in associated transmissibility. The *ahpC* mutation was not associated with an increased time-scaled haplotype density (*ahpC* mutants, median 0.19, IQR: 0.09- 0.22; *ahpC* non-mutants, median: 0.56, IQR: - 0.44-0.71, p = 0.93). Further, individuals with an incarceration history (currently or formerly incarcerated) were no more likely to be infected with a *M. tuberculosis* isolate with the *ahpC* mutation than were individuals with no incarceration history (*χ*2(1)=0.25, *p*=0.62).

## Discussion

We generated, to our knowledge, the first genomic portrait of circulating *M. tuberculosis* diversity and transmission dynamics to directly inform Paraguay’s national TB control program priorities. We found the majority of TB cases included in our study were attributable to recent transmission, including elevated rates of recent transmission within prisons and transmission clusters spanning prisons and surrounding communities that have expanded over the past twenty years. While *M. tuberculosis* is geographically structured in Paraguay, we identified a signal of continuous spread of *M. tuberculosis* between Paraguay’s major urban centers.

We found that most sampled infections were likely attributable to recent transmission rather than long-distance migration or activation of latent disease, similar to what has been reported in other medium-incidence countries^14^. Consistent with expectations that clustering rates may correlate with incidence, when applying a 5-SNP threshold, we found that isolates from Paraguay were more frequently clustered (45%) than those from a low-incidence setting in Spain (23%) and less frequently clustered than in a high-incidence setting in Mozambique (58%). Interestingly, we found a higher rates of clustering compared to what was reported in Malawi (36%), a high-incidence setting^11^. This could reflect the shorter, one year sampling timeframe of the Malawi study^11^, resulting in different genomic sampling rates, the use of different genomic sequencing pipelines, or true differences in transmission.

Paraguay’s incarceration rate has dramatically increased, from 60 per 100,000 people in 2000 to 194 per 100,000 in 2020^3,6,56^. More than seventy percent of the incarcerated population are pre-trial detainees, the highest proportion in South America^3^. The inhumane and unhealthy conditions of prison environments put people at heightened risk of disease and mortality; this risk translates into an increasing concentration of TB within prisons, with 18% (537/2,593) of notified TB cases in Paraguay occurring among incarcerated individuals in 2020^6^. Paraguay’s TB Control Program has worked in prisons since 2004 to provide trainings for healthcare providers and all diagnostic and treatment supplies, including laboratory capacity for microbiological testing in four prisons.

Our findings highlight the critical need to expand and strengthen existing programs to detect and treat TB early and to expand awareness and knowledge of the risks associated with prison environments. Isolates sampled from prisons were more frequently found in genomic transmission clusters and had a higher time-scaled haplotype density than did isolates from outside prisons, phylogenetic evidence that recent transmission was more frequent in prisons than in communities outside prisons. Further, *M. tuberculosis* sampled from prisons and the community were closely evolutionarily related and the majority of putative transmission clusters including individuals who were never incarcerated also included people who had a recent incarceration history, indicating that reducing transmission risk within prisons is an urgent public health priority with consequences both within and outside prisons.

While rates of drug resistance were relatively low, we found several phylogenetically unique mutations associated with both isoniazid and rifampicin resistance. These unique mutations could reflect either the *de novo* acquisition of a resistance mutation or the importation of a resistance mutation from outside Paraguay. Regardless, there is a critical need for expanding drug-susceptibility testing including both rapid testing for rifampicin resistance in addition to isoniazid monoresistance are critical to ensure patients are put on correct treatment courses and to reduce the risk of further resistance acquisition^19,20^.

The emergence of an independent *ahpC* mutation within a single sublineage opens questions about its phenotypic consequences. Previous studies in laboratory strains have reported that *ahpC* mutations are compensatory in the context of *katG* isoniazid resistance-conferring mutations, by recovering the bacterium’s ability to detoxify organic peroxides, but did not find measurable isoniazid resistance conferred by independent *ahpC* mutations^55^. Genome-wide association studies of clinical *M. tuberculosis* isolates confirmed the compensatory role of *ahpC* mutations^57^. *ahpC* mutations did not meet the criteria for being included in the 2021 WHO catalogue of resistance-conferring mutations because they were either too rare or had a low positive predictive value for isoniazid resistance as an independent mutation^35^. However, a study of isoniazid resistant isolates from Brazil reported that while *ahpC* mutations often co-occurred with *katG* mutations, they were also found in the absence of known resistance mutations in *katG* or *inhA*^58^.

A previous genome-wide survival analysis identified lineages and specific mutations associated with pre-resistance, genomic backgrounds that had a heightened likelihood of acquiring resistance-conferring mutations^59^. Whether *ahpC* acts as in a similar way, generating a “pre-compensated” genomic background, increasing the likelihood of future *katG* mutations, remains unknown.

Our study has several limitations. First, while we sequenced all available *M. tuberculosis* cultures, our final sample size of *M. tuberculosis* genomes was small relative to the number of notified TB cases in our study departments over the study period. Some locally circulating genotypes may therefore not be included in our sample and may lead to an underestimate of the contribution of recent transmission to incident TB. However, we sampled over a moderately long timeframe (five years) and included samples from high-incidence prisons and neighboring communities, providing greater opportunity to recover transmission events. Second, surveillance focused on Paraguay’s urban centers, where the majority of TB notifications occur. Future *M. tuberculosis* genomic surveillance in the Chaco, western Paraguay, where incidence is three times higher than in eastern Paraguay^15^, is needed.

Additionally, further analysis at the regional-level will be critical for understanding transmission between Paraguay and neighboring countries. Third, we sampled TB infections from prisons at a higher rate than infections outside prisons, potentially biasing upwards estimates of the rate of genomic clustering within prisons compared to outside prisons. Finally, we sequenced isolates from cultured sputum, as is routinely done for *M. tuberculosis* genomic epidemiology, but which limits the within-host diversity recovered from an individual’s infection. Future research is needed to develop sequencing approaches to recover within-host *M. tuberculosis* variation and incorporate this level of variation into transmission and ancestral state reconstruction.

Together, our results underscore an urgent need for TB control measures to interrupt ongoing transmission in Paraguay, particularly in high-incidence prison settings, which have an outsized role in broader transmission. Further, the connectivity of Paraguay’s urban centers indicates that TB control needs to be coordinated country-wide.

## Data Availability

Raw sequence reads for samples passing filters are available at the Sequence Read Archive (PRJNA870648).

## Acknowledgments

The authors thank the following health staff who facilitated the field work laboratory analyses and clinical follow up: Ruth Martinez, Nestor Moreno, Natalia Sosa. We also would like to express our gratitude to Pilar Muñoz, Johana Monteserin, CEDIC, LCSP and the Penitentiary Health Department from the Ministry of Justice for its support and advice for this project. The study was supported by the grant PIN15-705 from National Commission of Science and Technology (CONACYT) of Paraguay.

## Supporting information captions

**Figure S1. Genomic *M. tuberculosis* surveillance in Paraguay**. Flowchart indicates the total number of notified cases of TB in Paraguay, from 2016 to 2021; TB cases in the three major urban departments of Paraguay; TB cases in urban departments stratified by incarceration status at the time of TB notification; and number of cases for which sequenced *M. tuberculosis* passed all quality filters.

**Figure S2. Longitudinal changes in sampled *M. tuberculosis* genomic diversity**. We compared genomic diversity in our study (2016-2021) with that sampled in the only previous genetic study of *M. tuberculosis* in Paraguay (Candia et al. 2003). Stacked bar plots indicate the proportion of samples falling in each clade. From left to right, panels indicate the total diversity sampled, samples from Asunción, samples from Ciudad del Este, and samples from the 2003 study. Panels from the current study are stratified by incarceration status at the time of TB notification; the 2003 study did not present data stratified by incarceration status.

**Figure S3. Time-scaled haplotype density by incarceration status at the time of TB notification**. We measured time-scaled haplotype diversity, a measure of the centrality of a single tip isolate to all other isolates on the tree, an alternative proxy for recent transmission that considers not only the nearest phylogenetic neighbor, but all tree trips. We calculated time-scaled haplotype density from a matrix of pairwise SNP distances with the R package *thd* as previously described^52^. We set the *M. tuberculosis* substitution rate to 1×10^−7^ substitutions per site per year and included an effective genome length of 3,916,441 basepairs (the length of the reference genome minus the PE/PPE regions excluded from variant calling) and used short (10 year) and long (20 year) epidemic timescales. Boxplots are colored by incarceration status at the time of TB notification. Boxes indicate the interquartile range, lines indicate median values, and whiskers indicate the range of the data. Points outside the whiskers indicate outliers.

