## Supplementary Information for "Phylogeography and transmission of *M. tuberculosis* spanning prisons and surrounding communities in Paraguay"

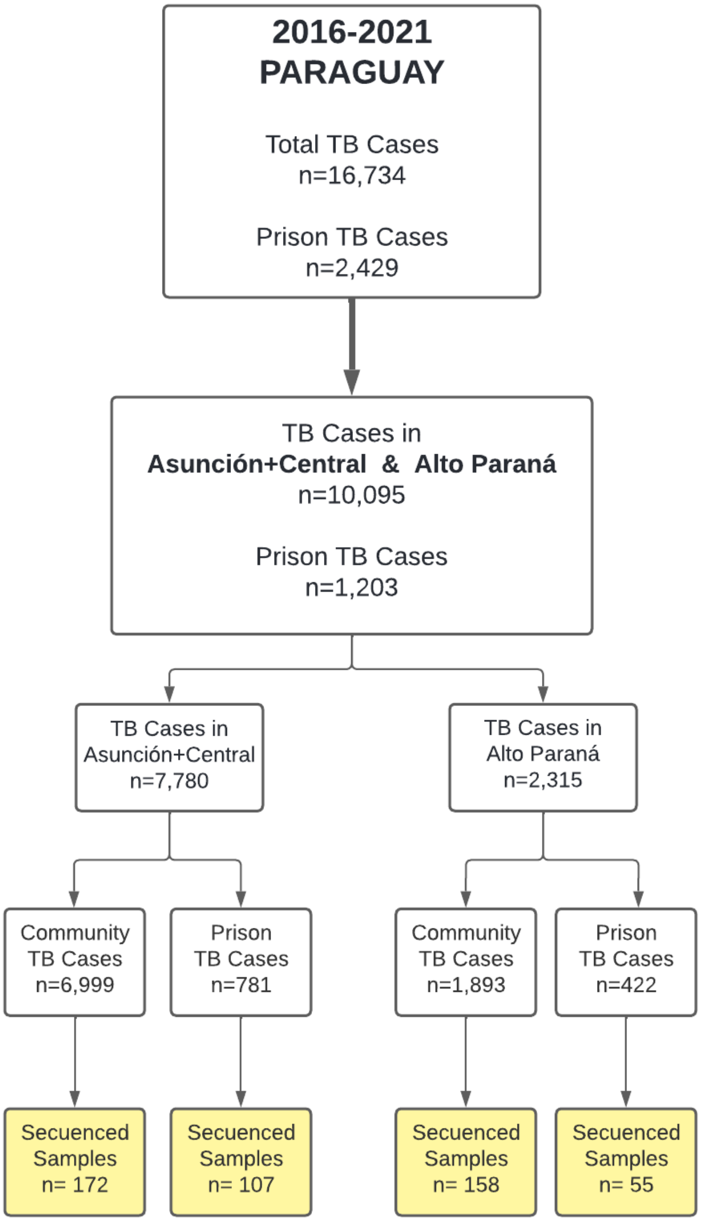


**Figure S1. Genomic *M. tuberculosis* surveillance in Paraguay.** Flowchart indicates the total number of notified cases of TB in Paraguay, from 2016 to 2021; TB cases in the three major urban departments of Paraguay; TB cases in urban departments stratified by incarceration status at the time of TB notification; and number of cases for which sequenced *M. tuberculosis* passed all quality filters.

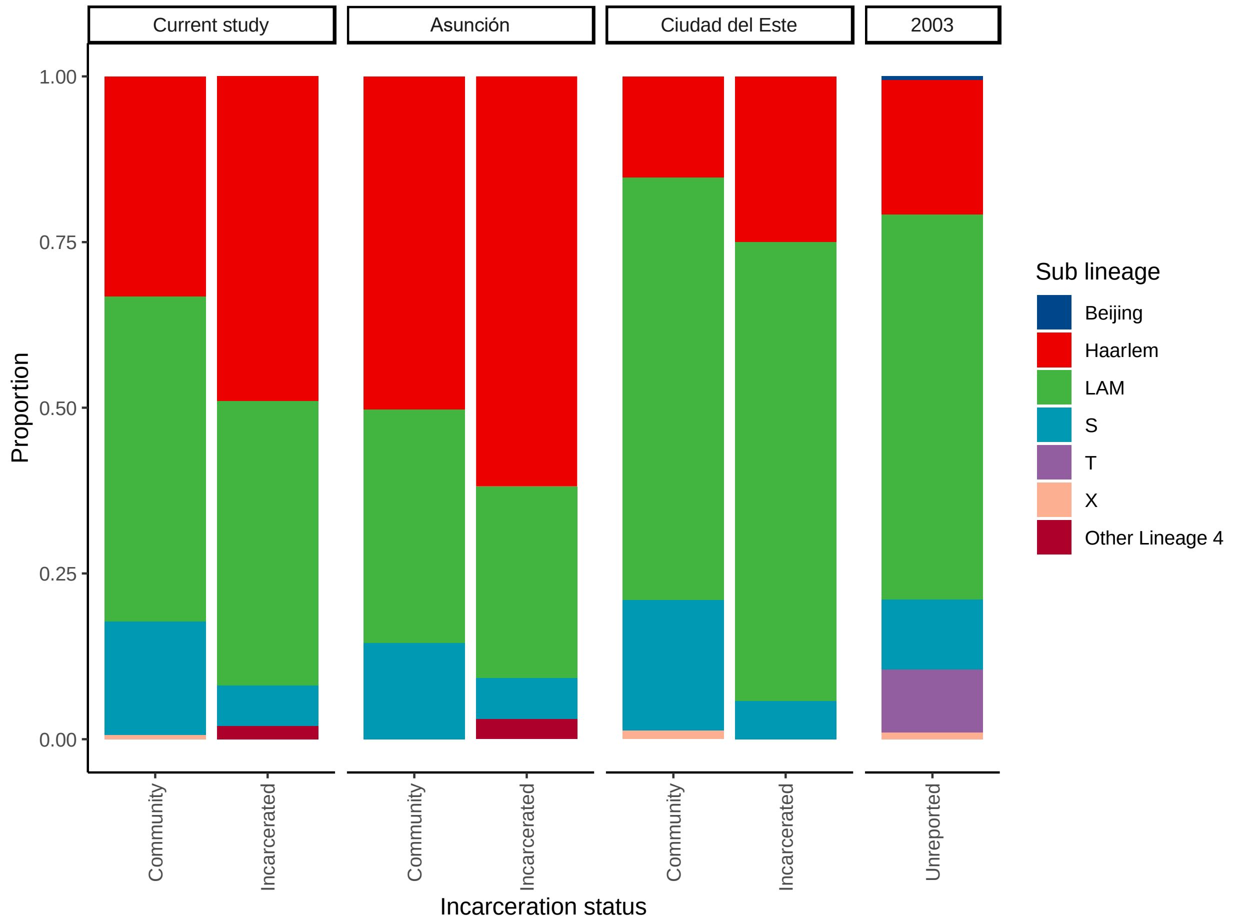


**Figure S2. Longitudinal changes in sampled *M. tuberculosis* genomic diversity.** We compared genomic diversity in our study (2016-2021) with that sampled in the only previous genetic study of *M. tuberculosis* in Paraguay (Candia et al. 2003). Stacked bar plots indicate the proportion of samples falling in each clade. From left to right, panels indicate the total diversity sampled, samples from Asunción, Ciudad del Este, and from the 2003 study. Panels from the current study are stratified by incarceration status at the time of TB notification; the 2003 study did not present data stratified by incarceration status.

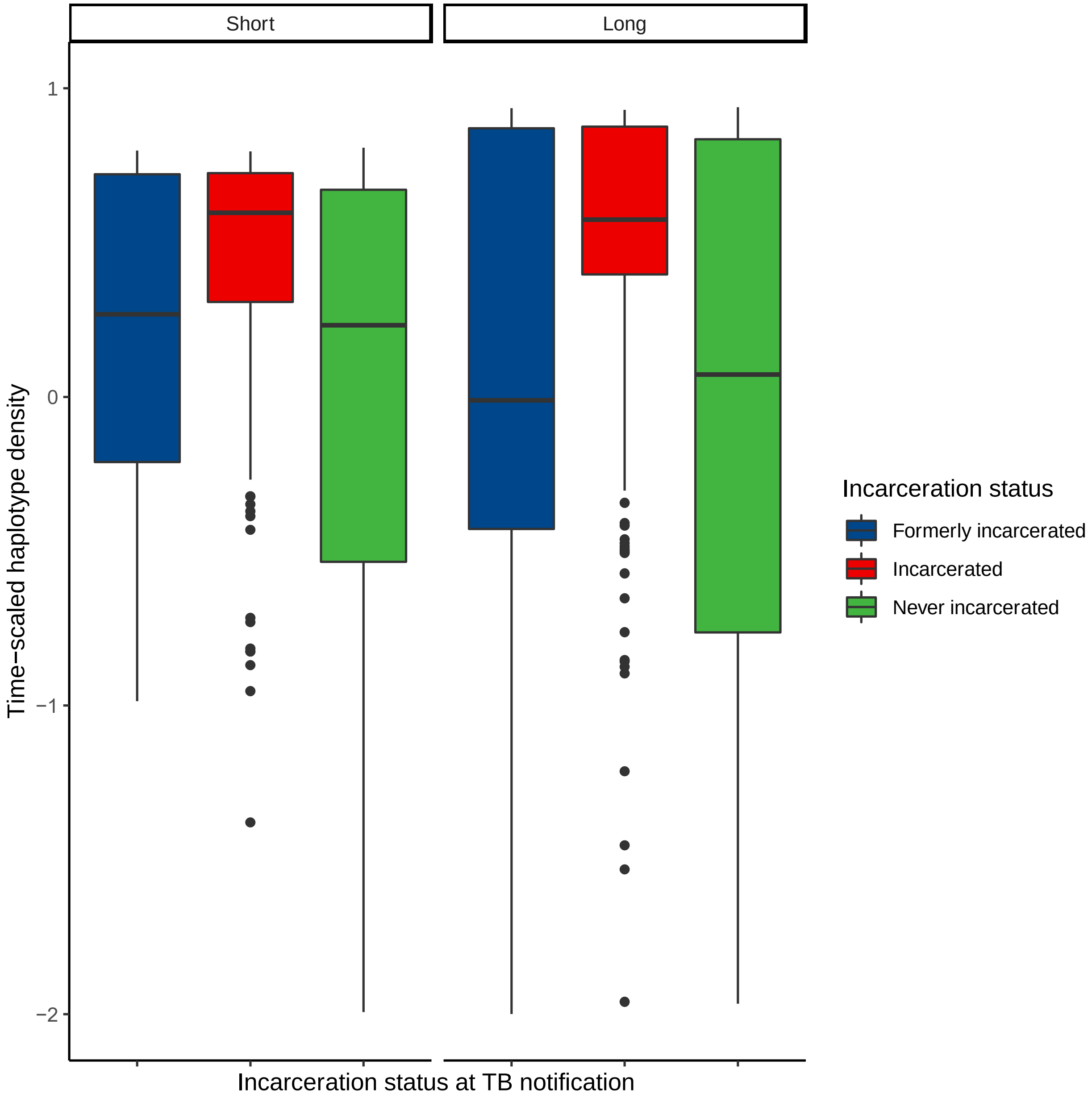


**Figure S3. Time-scaled haplotype density by incarceration status at the time of TB notification.** We measured time-scaled haplotype diversity, a measure of the centrality of a single tip isolate to all other isolates on the tree, a proxy for recent transmission that considers not only the nearest phylogenetic neighbor, but all tree trips. We calculated time-scaled haplotype density from a matrix of pairwise SNP distances with the R package *thd* as previously described[53]. We set the *M. tuberculosis* substitution rate to $1x10^{-7}$ substitutions per site per year and included an effective genome length of 3,916,441 basepairs (the length of the reference genome minus the PE/PPE regions excluded from variant calling) and used short (10 year) and long (20 year) epidemic timescales. Boxplots are colored by incarceration status at the time of TB notification. Boxes indicate the interquartile range, lines indicate median values, and whiskers indicate the range of the data. Points outside the whiskers indicate outliers.
